# Automated Contouring and Planning for Hippocampal-Avoidant Whole-Brain Radiotherapy

**DOI:** 10.1101/2020.04.20.20072827

**Authors:** Christine H. Feng, Mariel Cornell, Kevin L. Moore, Roshan Karunamuni, Tyler M. Seibert

## Abstract

**Purpose:** Design and evaluate a workflow using commercially available artificial intelligence tools for automated hippocampal segmentation and treatment planning to efficiently generate clinically acceptable hippocampal-avoidant whole brain (HA-WBRT) radiotherapy plans.

**Methods and Materials:** We retrospectively identified 100 consecutive adult patients treated for brain metastases outside the hippocampal region. Each patient’s T1 post-contrast brain MRI was processed using FDA-approved software that provides segmentations of brain structures in 5-7 minutes. Automated hippocampal segmentations were reviewed for accuracy and edited manually if necessary, then converted to files compatible with a commercial treatment planning system, where hippocampal avoidance regions and planning target volumes (PTV) were generated. Other organs-at-risk (OARs) were previously contoured per clinical routine. A RapidPlan knowledge-based planning routine was applied for a prescription of 30 Gy in 10 fractions using volumetric modulated arc therapy (VMAT) delivery. Plans were evaluated based on NRG CC001 dose-volume objectives.

**Results:** Of the 100 cases, 99 (99%) had acceptable automated hippocampi segmentations without manual intervention. Knowledge-based planning was applied to all cases; the median processing time was 9 minutes 59 seconds (range 6:53 – 13:31). All plans met per-protocol dose-volume objectives for PTV per the NRG CC001 protocol. For comparison, only 66.0% of plans on NRG CC001 met PTV goals per protocol, with 26.3% within acceptable variation. In this study, 43 plans (43%) met OAR constraints, and the remaining 57 (57%) were within acceptable variation, compared to 42.9% and 48.6% on NRG CC001, respectively. No plans in this study had unacceptable dose to OARs, compared to 0.8% of manually generated plans from NRG CC001.

**Conclusion:** An automated pipeline harnessing the efficiency of commercially available artificial intelligence tools can generate clinically acceptable VMAT HA-WBRT plans with minimal manual intervention. This process could improve clinical efficiency for a treatment established to improve patient outcomes over standard WBRT.

## Introduction

Whole-brain radiotherapy (WBRT) remains an important treatment for patients with multiple brain metastases, with over 200,000 cancer patients treated with WBRT in the United States annually^1^. Compared to stereotactic radiosurgery (SRS), WBRT provides better distant intracranial tumor control, at a cost of decreased control of existing intracranial metastases and increased neurocognitive adverse effects^2,3^. Hippocampal-avoidant WBRT (HA-WBRT) has emerged as an approach to retain the intracranial tumor control of WBRT while minimizing cognitive decline.

Neurocognitive dysfunction following irradiation can occur through depletion of hippocampal neural stem cells as they differentiate to a gliogenic lineage and hippocampal atrophy^4^. Hippocampal dosimetry is associated with long-term decline in list-learning delayed recall^5^. A multi-institutional phase II trial, RTOG 0933, demonstrated that hippocampal-avoidant WBRT (HA-WBRT) provided improved preservation of memory and quality of life compared to historical controls^6^. More recently, NRG CC001, a phase III trial that randomized patients to standard WBRT with memantine or HA-WBRT with memantine, demonstrated better cognitive preservation and quality of life without difference in intracranial tumor control or overall survival^7^.

The neurocognitive and quality of life advantages of HA-WBRT over standard WBRT provide a compelling argument for HA-WBRT to be considered the new standard of care for patients with good performance status who will undergo WBRT. However, manual hippocampal contouring and IMRT planning are significantly more complex and time-consuming than the blocks and 3D conformal planning of traditional WBRT^8,9^. We designed and evaluated a workflow using commercially available artificial intelligence tools for automated hippocampal segmentation and treatment planning to efficiently generate clinically acceptable HA-WBRT plans.

## Methods and Materials

### Study Design & Patients

We retrospectively identified 100 consecutive adult patients who received radiotherapy for brain metastases at UC San Diego between April 2015 – August 2018. Eligible patients had an available brain MRI showing intracranial metastases no closer than 5 mm from the hippocampus. Per clinical routine, all of these brain MRI volumes already had associated contours for the brain and for standard organs-at-risk (OARs), including bilateral lens, bilateral optic nerves, and optic chiasm. This study was reviewed and approved by the UC San Diego Institutional Review Board (IRB #181609).

### Hippocampal Segmentation

Thin-slice T1 brain MRIs were processed using NeuroQuant (CorTechs Labs, Inc., San Diego, CA, USA), an FDA-approved software. Our segmentations were performed as untimed batch processing jobs for convenience. The vendor states that segmentations of bilateral hippocampi are typically generated in 5-7 minutes per patient or MRI volume^10,11^. Outputs from NeuroQuant were converted to RTSTRUCT DICOM files compatible with a commercial treatment planning system, Eclipse version 15.6 (Varian Medical Systems, Palo Alto, CA, USA), using software developed in-house with Matlab (Mathworks, Natick, MA, USA). Automated hippocampal segmentations were reviewed for accuracy by a radiation oncologist and edited manually, if necessary. The number of patients requiring manual edits was recorded.

### Knowledge-Based Planning

The imported segmentations and MRI were automatically registered to the patient’s simulation CT using Eclipse. Hippocampal avoidance regions were generated using 5 mm uniform expansion, and planning target volumes (PTV) were generated by subtracting the hippocampal avoidance region from the existing brain contour. Standard OARs (bilateral lens, bilateral optic nerves, and optic chiasm) were previously contoured per clinical routine. A publicly available RapidPlan (Varian Medical Systems, Palo Alto, CA, USA) knowledge-based planning routine^12^ was applied for a prescription of 30 Gy in 10 fractions using volumetric modulated arc therapy (VMAT) delivery via four full arcs on a TrueBeam (Varian Medical Systems, Palo Alto, CA, USA) linear accelerator with 120-leaf Millennium multileaf collimators. The processing time, defined as the time from image registration to completion of dose calculation, was recorded using a stopwatch for all cases.

### Plan Evaluation

Plans were normalized to deliver prescription dose to 95% of the PTV. Dose-volume histograms (DVH) were calculated for the PTV and OARs of each plan. Results were evaluated based on NRG CC001 dose-volume objectives (Table 2).

## Results

A summary of scan characteristics is in Table 1. The scans were split approximately evenly between 1.5 tesla and 3.0 tesla systems. Sixteen (16%) cases had a resection cavity on the scan. Of the 100 cases, 99 (99%) had acceptable automated hippocampi segmentations, without any manual intervention (Figure 1). One case required minor manual editing at the junction of the hippocampus and the lateral ventricle due to the hippocampus segmentation extending 6 mm into the lateral ventricle. In comparison, on central review of the OARs used for participants on NRG CC001, only 66% of OAR contours were per protocol.

**Table 1.** Summary of MRI characteristics for all cases.

|  |  | N (%) |
| --- | --- | --- |
| **Magnet Strength (T)** | 1.5 | 53 (53%) |
|  | 3 | 47 (47%) |
| **Slice Thickness (mm)** | 1 | 94 (94%) |
|  | 1.2 | 1 (1%) |
|  | 1.5 | 1 (1%) |
|  | 2 | 4 (4%) |
| **IV Contrast** | Yes | 98 (98%) |
|  | No | 2 (2%) |
| **Scanner Model** | GE SignaHDxt 1.5T | 53 (53%) |
|  | GE SignaHDxt 3.0T | 28 (28%) |
|  | GE Discovery MR750 3.0T | 10 (10%) |
|  | GE Discovery MR750w 3.0T | 9 (9%) |
| **Resection Cavity** | Yes | 16 (16%) |
|  | No | 84 (84%) |

**Figure 1.**
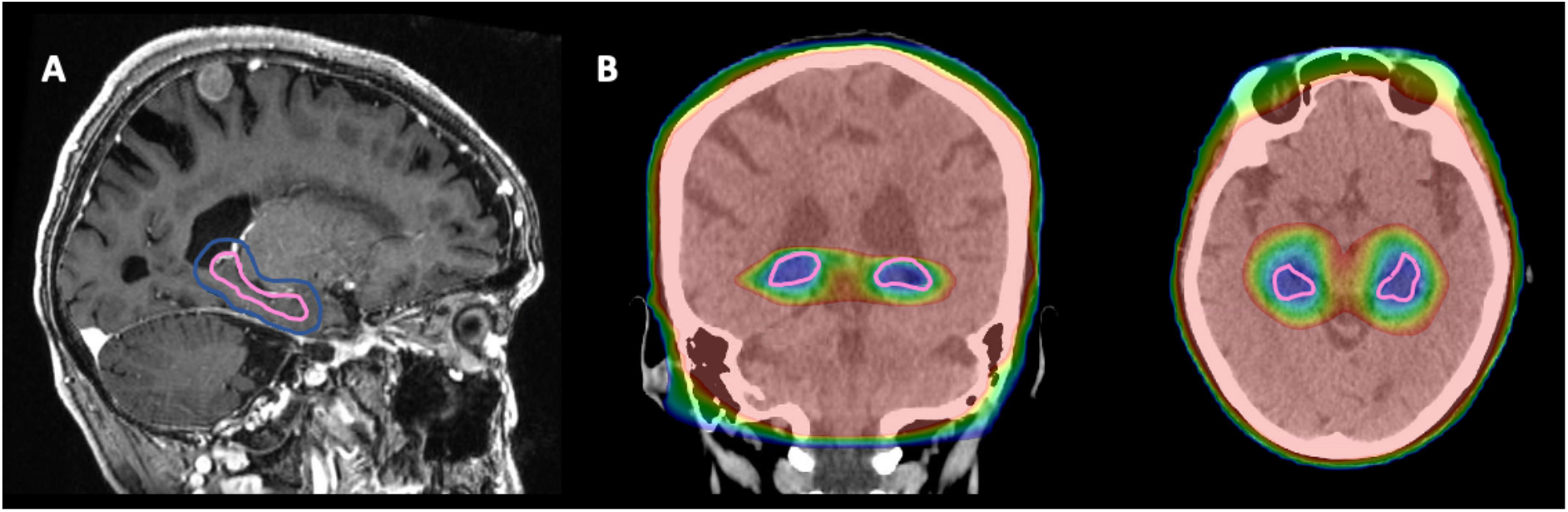
Representative case with A) processed hippocampi segmentation (pink) and hippocampal avoidance region (dark blue), B) dose in color wash ranging from 8 Gy (blue) to 30 Gy (red).

Knowledge-based planning was applied to all cases, with individual optimization settings. Eleven (11%) cases were processed by an experienced dosimetrist. Eighty-nine (89%) cases were processed by a radiation oncologist after undergoing a 20-minute training session with a dosimetrist. The median processing time for all plans was 9 minutes 59 seconds (range 6 min 53 sec – 13 min 31 sec). There was no difference in planning time between the dosimetrist and the radiation oncologist.

All plans met acceptable dose-volume objectives for PTV and OARs per the NRG CC001 protocol (Table 2). The case that required manual editing was planned using the manual contours, then again using the automatically segmented contours for PTV and PRV delineation. Both plans met per-protocol dose-volume objectives for the corrected PTV and OARs. PTV doses were per protocol for all plans, with D_2%_ below 37.5 Gy and D_98%_ greater than 25 Gy. For comparison, only 66.0% of plans on NRG CC001 met PTV goals per protocol, with 26.3% within acceptable variation^7^.

**Table 2.** Mean DVH metrics for HA-WBRT in current study compared to NRG CC001 constraints.

|  | NRG CC001 |  |  |
| --- | --- | --- | --- |
|  | Per Protocol | Acceptable Variation | Current study Mean (range) |
| **PTV D<sub>2%</sub> (GY)** | ≤ 37.5 | 37.5-40 | 34.3 (33.3-37.2) |
| **PTV D<sub>98%</sub> (GY)** | ≥ 25 | 22.5-25 | 26.5 (25.3-28.8) |
| **PTV V<sub>30GY</sub> (%)** | ≥ 95 | 90-95 | 95 (95-95) |
| **HIPPOCAMPUS D<sub>100%</sub> (GY)** | ≤ 9 | 9-10 | 8.1 (7.4-9.1) |
| **HIPPOCAMPUS D<sub>MAX</sub> (GY)** | ≤ 16 | 16-17 | 13.1 (9.7-15.7) |
| **OPTIC NERVES D<sub>MAX</sub> (GY)** | ≤ 30 | 30-37.5 | 29.8 (25.4-32.3) |
| **OPTIC CHIASM D<sub>MAX</sub> (GY)** | ≤ 30 | 30-37.5 | 30.1 (29.0-32.0) |

Abbreviations: D, dose; PTV, planning target volume.

Dose to bilateral hippocampi was per protocol for 99 plans (99%), with one plan delivering D_100%_ of 9.05 Gy (per protocol was D_100%_ ≤ 9.0 Gy, with 9-10 Gy acceptable variation). Hippocampal D_max_ was less than the per protocol recommendation of 16 Gy for all plans. Forty-three plans (43%) met OAR constraints for optic structures, and the remaining 57 plans (57%) were within acceptable variation. The highest D_max_ for any optic structure across all plans was 32.3 Gy, well below the protocol maximum of 37.5 Gy. On NRG CC001, 42.9% of plans met OAR constraints per protocol, and 48.6% were within acceptable variation^7^. No plans in this study had unacceptable dose to OARs, compared to 0.8% of manually generated plans from NRG CC001.

## Discussion

Randomized clinical trial results have established HA-WBRT as superior to WBRT for preservation of neurocognitive function and quality of life^6,7^. Using commercially available software for hippocampal contours and knowledge-based planning, we have established a workflow to generate automated HA-WBRT plans with meaningful efficiency. Standard clinical MRI data were used from either 1.5 or 3.0 tesla systems. Automated hippocampal volumes were accurate without any manual intervention in 99% of cases. Knowledge-based planning typically required approximately 10 minutes and yielded HA-WBRT plans that were more frequently adherent to the NRG CC001 protocol than the manually generated plans actually used in that trial^7^.

Hippocampal-avoidant WBRT gained attention after publication of the results from RTOG 0933, and the recent results from NRG CC001 demonstrate improved patient outcomes over standard WBRT. Particularly as cancer therapies have improved survival rates, preservation of neurocognitive function and quality of life is increasingly important. However, manual hippocampal contouring can be challenging with high interobserver variability^13,14^, and inverse plan optimization can require multiple iterations to fulfill constraints with generation of helping structures. We have shown that automated tools could be integrated into this complex process to improve both efficiency and plan quality.

The knowledge-based plans provided similar dose distributions to previously published manual and automated planning studies^8,15–17^. In the studies from Gondi et al. and Nevelsky et al., plans were created using nine linac-based IMRT fields, while Krayenbuehl et al. used four non-coplanar arcs and Wang et al. studied both IMRT and VMAT approaches^8,15–17^. Our plan metrics differ most notably in improved PTV coverage at the expense of slightly higher hot spots, compared to Krayenbuehl et al.^15^, and decreased maximum dose to optic structures, in relation to the other studies^16,17^. The automated workflow presented here also demonstrated a consistent plan quality across cases, with none exceeding protocol constraints.

One criticism of typical WBRT is poor local control of existing brain metastases after 30 Gy. A recent single-arm feasibility study investigated the utility of a simultaneous integrated boost (SIB) technique with WBRT^18^. The investigators prescribed 30 Gy in 12 fractions to the brain and simultaneously boosted metastases and resection cavities to 42 or 51 Gy. Comparison to propensity matched patients treated with conventional WBRT demonstrated increased intracranial progression-free survival and overall survival with HA-WBRT with SIB. The ongoing HIPPORAD trial (NOA-14, ARO 2015-3, DRKS00004598) will further study this method. If that approach is successful, further development of the workflow presented here could incorporate SIB technique into knowledge-based planning.

A limitation of this study is that time for OAR contours was not included in the planning time measurement. The hippocampal contours were generated automatically (except for minor manual edits in 1% of cases); although not timed here, these contours are typically generated in 5-7 minutes, per the software vendor^11^. Standard OARs (brain, optic nerves, lenses, and optic chiasm) had been contoured previously and were also not timed. Future investigations could include automated contours of these standard structures, which are generally familiar and routinely included in most brain radiotherapy plans. The inclusion of automatically contoured standard OARs into this process would make the system completely autonomous, i.e. a class solution pipeline that takes as input standard imaging and outputs a complete HA-WBRT plan with no human-driven parameters.

In conclusion, an automated pipeline harnessing the efficiency of commercially available artificial intelligence tools can consistently generate clinically acceptable VMAT HA-WBRT plans with minimal manual intervention. This process could improve clinical efficiency for a treatment established to improve patient outcomes over standard WBRT.

## Data Availability

The data that support the findings of this study are available from the corresponding author upon reasonable request.

## Acknowledgements

None

